# Feedback of Individual Genetic and Genomics Research Results: A Qualitative Study Involving Grassroots Communities in Uganda

**DOI:** 10.1101/2022.04.08.22273613

**Authors:** Joseph Ochieng, Betty Kwagala, John Barugahre, Marlo Möller, Keymanthri Moodley

## Abstract

**Background:** Genetics and genomics research (GGR) is associated with several challenges including, but not limited to, implications of sharing research findings with participants and their family members, issues of confidentiality, determining appropriate methods for providing genetic or genomic information to individuals tested, and ownership of DNA obtained from the samples. Additionally, GGR holds significant potential risk for social and psychological harms.

A considerable amount of research has been conducted with resultant literature and global debate on return of genetic and genomics testing results, but such investigations are limited in the African setting, including Uganda.

The objective of the study was to assess perceptions of grassroots communities on if and how feedback of individual genetics and genomics testing results should be carried out in a Ugandan setting.

**Methods:** This was a cross-sectional study that employed a qualitative exploratory approach. A total of 42 individuals from grassroots communities representing three major ethnic groupings participated in five deliberative focus group discussions. Data were analysed through content analysis along the main themes of the study. NVivo software (QSR international 2020) was used to support data analysis and illustrative quotes were extracted.

**Results:** Of the 42 respondents 23 (55%) were male with an age range of 18-77 years. Most (70%) were small scale farmers, and the majority were Christians, who were married and had children. They all lived in a rural community in one of the three regions of the country and had no prior participation in GGR. All the respondents were willing to undergo genetics testing and receive feedback of results with the main motivation being diagnostic and therapeutic benefits as well as facilitating future health planning. Content analysis identified three themes and several sub-themes including 1) the need to know one’ s health status; 2) ethical considerations for feedback of findings and 3) extending feedback of genetics findings to family and community

**Conclusion:** Participation in hypothetical genetics and genomics research as well as feedback of testing results is acceptable to individuals in grassroots communities. The strong therapeutic misconception linked to GGR is concerning and has implications for consent processes and genetic counselling. Privacy and confidentiality, benefits, risks as well as implications for sharing need to be considered for such feedback of results to be conducted appropriately.

## Introduction

Although the expanding applicability of knowledge generated from genetics and genomics research (GGR) holds great promise for discoveries in the biomedical and socio-behavioural sciences, it also raises challenging ethical and societal issues. Such challenges include, but are not limited to, implications of sharing research findings with participants and their family members, issues of confidentiality, determining appropriate strategies for providing genetic or genomic information to individuals tested, and ownership of Deoxyribonucleic Acid (DNA) obtained from the samples [1-3]. Furthermore, GGR has significant potential risk for social and psychological harms, for example, studies that generate information about an individual’ s health risks can provoke anxiety and confusion, damage familial relationships, and/or compromise the individual’ s future financial status [4-7]. Results could also possibly be used as a basis for ethnic/racial segregation or discrimination such as denial of insurance coverage or employment [8].

A significant amount of research has been conducted with resultant literature and global debate on return of genetic and genomics testing results [9-14]. Despite the fact that international policies for return of individual genetic research findings are still evolving, general consensus appears to be that in order to consider findings for feedback a number of criteria need to be met including the ability to assess the evidence base for potentially disease causing genetic variants in relation to the concerned population(s); assessment of whether the particular finding is beneficial to the individual; ensuring that patients are appropriately informed of the implications of the findings for their disease or treatment, and referral for follow-up care while seeking guidance of the Research Ethics Committee (REC) [15]. However, such debate with a focus on the issues that affect the African setting is still limited [16-22]. This situation is exacerbated by the fact that many countries in the African region lack ethical guidelines on how such ethical issues can be addressed [23]. GGR has been conducted for about 20 years in the Ugandan setting and is expected to continue to increase owing to its potential for advancing targeted disease detection and interventions for both communicable and non-communicable diseases in this resource-limited setting [24]. Yet there is a paucity of knowledge on the ethical, legal and social challenges that accompany GGR in the country [25-28]. There have been a few publications on perspectives of researchers [26, 28] and research participants but virtually no published literature on perspectives of grassroots communities who are based in rural settings and are considered to have limited interaction with the outside world.

We set out to assess grassroots communities’ perceptions on whether feedback of individual genetics and genomics testing results should occur in a Ugandan setting to inform research ethics guideline development.

## Methods

### Study design and Setting

This was a cross-sectional study that employed a qualitative exploratory approach. The study was conducted by a team of academics comprising bioethicists and medical scientists with experience in qualitative research. JO a male medical doctor and academic with bioethics training and experience, BK a female PhD sociology academic of more than 20 years and JB a male PhD Philosophy academic led most of the interviews. They were assisted by eight research assistants including four females proficient in local languages. Data was collected between January and February 2021. Participants were recruited from remote grassroots communities in three regions of Uganda each representing a major ethnic grouping. Five deliberative focus group discussions (dFGDs) involving 42 participants were conducted across the three regions of the country. Two dFGDs were conducted in each region with one involving youth 18-35 years and the other involving individuals of 36 years and above. However, in one of the regions only one FGD involving individuals older than 35 years was conducted. The communities were selected from the eastern, northern and west Nile regions of Uganda to represent the main ethnic groupings. Participants were recruited from predetermined ethnic groups, districts and sub-counties. The specific local communities were selected by the research assistants identified at the respective sub-counties.

### Data collection

The dFGDs were conducted in open spaces in the compounds of health facilities, schools or churches a safe distance away from non-participants. Data was collected in accordance with the Covid-19 prevention measures including hand sanitization, face masking, social distancing and in open spaces of compounds under trees in order to limit any potential for infection spread

Data collection entailed face to face deliberative focus group discussions lasting between 90 to 120 minutes and were conducted in the respective local languages of the concerned community. Initially, participants were asked general questions on awareness and knowledge about genetics and genomics. This was followed by a 30-minute explanatory session on the meaning and role of genetics and genomics as well as the testing and feedback of results lasting about 30 minutes. This education session was followed by a discussion moderated by the FGD guide. The discussion included willingness to participate in GGR, willingness to receive feedback following genetic testing, conditions for feedback and extending feedback to family and community. The discussions were audio recorded and complemented by notes taken by a research assistant.

### Data management and analysis

Recorded information was transcribed verbatim, checked for accuracy and later translated into English. Data were analysed through content analysis along the main themes of the study. Content analysis was conducted using a comprehensive thematic matrix that included identifying codes, categories and themes to identify common patterns arising from the narratives. The coding was done both deductively and inductively, whereby we started deductively with a set of codes, but then inductively come up with new codes as we sifted through the data. Transcripts were further reviewed for emerging themes which were integrated into the thematic matrix. The researcher, JO was involved in applying and confirming application of codes across all transcripts and disagreements were resolved by cross checking with the recorded data. NVivo software (QSR international 2020) was used to support data analysis and illustrative quotes were extracted.

### Ethical considerations

Ethical review and approval was obtained from the Makerere University School of Biomedical Sciences Higher Degrees and Research Ethics Committee ref. SBS 628 and the Health Research Ethics Committee, Stellenbosch University ref. HREC 16853, followed by clearance by the Uganda National Council for Science and Technology (UNCST) ref. SS268ES.

Both male and female individuals of 18 years and above who had provided written informed consent participated in the study. No participant identifying information was recorded.

### Findings

Of the 42 respondents 23 (55%) were male, age range 18-77 years, 70% small scale farmers, and majority were Christians, married and had children. They all lived in a rural community in one of the three regions of Uganda and had no prior genetics and genomics research experience. All the respondents were willing to undergo genetics testing and receive feedback of results. The main reasons for receiving results were the need to know one’ s health condition and to seek care or plan for the future as well as that of their loved ones.

Content analysis identified three themes and a number of sub-themes including 1) the need to know one’ s health condition, with subthemes of benefits of feedback and concerns, challenges and implications for sharing; 2) considerations for feedback of findings, with sub-themes adequate informed consent, genetic counselling as well as privacy and confidentiality; and 3) extending feedback of genetics findings to family and community.

#### 1. The need to know one’ s health condition

Almost all the respondents replied in the affirmative when asked about their willingness to receive feedback of hypothetical genetic test results because they felt that it was useless to take the test if they would not receive results. All respondents stated that genetic testing is acceptable and would contribute to improved knowledge of the field. All respondents also indicated that findings of such testing need to be shared with individuals tested because it was considered important to know one’ s health status. Knowing of one’ s genetics information was a major motivating factor for participating in genetics and genomics research or taking a genetic test.

> *“When I go to test, I go because I know that I want to know my health status, so if am tested and they don’ t give me my results it is almost like I have not done any tests, so if I get tested my results should be brought back so that in case I have any underlying conditions I can look for help*.*’ ‘* ***FGD 007***
>
> *“I want to know my results because if you test for anything you have to know your results so that I can know if am healthy or sick*.*”* **FGD 008**
>
> *“Yes, it should be given. It should be given to me the patient so that I can know exactly what they have found out*.*’ ‘* **FGD 007**

Respondents felt that for any test carried out, the results will either turn out to be positive or negative, and for any underlying condition the results will turn out positive meaning that feedback helps one to start living a new life. Respondents noted that even if the treatment may not be available for the diagnosed condition, it would still help them understand their health condition and plan for their future. Thus, if results are not shared, the individuals tested will remain unsettled and anxious wondering what could be happening to their bodies. Some thought that if treatment is not available at the testing centre, it could as well be sought from other hospitals provided one knew what their health problem is.

> *“I want to know the results of those tests because I want to know my health status so that if am sick, I go to the hospital, if am not I start planning my life afresh*.*’ ‘* ***FGD 008***
>
> *“It is right because it helps me to know my status which gives me the strength to take care of my children*.*’ ‘* ***FGD 008***

A minority of respondents felt that they would wish to know their test results only if the condition is treatable. Otherwise, it would be stressful and cause unnecessary anxiety for one to be told of a disease, yet it has no available treatment.

‘*‘Using my body parts, I am not interested. If they are to teach me my blood group, I understand but if it is something else am not interested*.*’ ‘* **FGD 007**

Respondents had various reasons for wanting to know the results of their hypothetical genetics and genomics testing including being able to plan for the future, knowing their health conditions and being able to resolve some of the community myths particularly following death of individuals. For others, since the samples were from their bodies, they had a right to know the outcomes of the testing and researchers are obligated to provide such feedback. Some felt that knowing the results would be helpful in guiding the individuals on seeking therapy early enough.

> “*So that people can clearly know the actual cause of the death of a person, not that they are left to imagine*.” **FGD009**
>
> *“The results of the DNA should be given to me because it was part of my body that was removed, it was nobody’ s body part, it was mine*.*’ ‘* ***FGD 007***
>
> *“I think the results should be given to you because by the time you went for the test you wanted to know your health status so the results should be given to you so that you can know about your health status better*.*’ ‘* ***FGD 007***

#### Benefits of feedback for genetics findings

Respondents highlighted several benefits associated with feedback of genetic testing results including the fact that it helps individuals to know what to do in their life and that of their relatives. It can guide the medical professionals and scientists to search for treatment, and institute preventive measures before disease manifests. It also facilitates the government to plan and build hospitals that will specialise in managing those diseases. Others felt that it will be an added advantage because they will have gained more health information about themselves to help predict the future.

> ‘*‘I think it is basically the knowledge after getting the information that really prepares you to be free. Now like us at least we have heard and we have gotten to know what it is all about, so it gives me the freedom (courage) to participate freely without the fear that I had before*.’’ **FGD 006**
>
> ‘*‘It would help me know what the illness is and whether the complication is from my mother or my father, so that I can alert them and see how to protect my children*.’’ **FGD009**
>
> ‘*‘When I receive feedback at the right time and there are no other discouragements and at the same time the person who is giving me feedback first begins by counselling and guiding me, reminding me of what went on and how to live afterwards*.’’ **FGD 006**

Others felt genetic testing and associated feedback of results is good because they get to know their health condition and plan on how to protect themselves in case, they have potential to develop any illness. Some thought it an added advantage because in certain circumstances individuals live in an environment of uncertainty and suspicion for particular traits, yet after testing you can confirm your genetic lineage which would explain why things are happening that way. A desire for ancestral information was expressed.

> *“We know that these diseases could have come from the ancestral line of our parents, so knowing the result is good because you can be able to trace whether it is coming from your mother’ s line or father’ s line and inform them to protect the next generation of the family*.*’ ‘* **FGD 009**
>
> *“Yes, I want to know the results of this DNA test because it helps me to know the status of my blood and also know my clan too*.*’ ‘* FGD 008

About one third of the respondents including both men and women linked participation in genetics and genomics research as well as receiving feedback of results to establishing the paternity of their children if other family members were tested too.

> *“I want to acknowledge that my father made my sister to go through the same. At first, he denied being the father to my sister but when they went for a DNA test, it was confirmed that he was the true father. He no longer has any doubts and he is instead happy now*.*’ ‘* ***FGD 006***
>
> *‘’ In fact, this has happened to me before; my husband denied my second child saying I cheated and when we went to the hospital to prove, their DNA was the same and he even did not apologize for accusing me of adultery’ ‘*. ***FGD 009***
>
> ‘*‘It is a good thing because there have been cases of domestic violence because of a man doubting the paternity of some of his children, such would help solve some of these problems causing violence in the homes*.*’*’ **FGD 009**

#### Concerns, Challenges and implications for feedback of genetics and genomics testing results

Respondents noted that although genetics and genomics research and testing as well as the associated feedback of results is good, they have experienced situations where disclosure of cancer results to patients was felt to have hastened death. They also noted that if one has a 50% chance of developing or not developing cancer, giving such information can cause some trauma, hence the need for caution. Others were worried about the cost of such testing which they expected to be too much for them to afford, hence appealed for affordable genetics testing costs within reach of the low-income earners.

> ‘*‘To bridge the gap of language barrier like when somebody is an illiterate and does not know how to read and write, if the research department was able to bring the projection in form of a video, somebody who does not know how to read will be able to interpret what is going on. So, it would influence that person and attract more attention to that*.*’ ‘* **FGD 006**
>
> Some respondents noted that feedback of genetics results has the potential to reveal discordance in paternity and this has the potential to cause family break ups and associated psychological harm and suffering both to the child and the discordant parent. There was also concern about being diagnosed with a condition that is beyond the affordability of the family which could end up consuming all the family resources.
>
> ‘*‘The problem is, it is a bit expensive and then those services are very far, otherwise I would really say that it is a good thing to do*.’’ **FGD 006**
>
> “*The DNA is what distinguishes one person from another. For example, you can tell that this child does not belong to this family and does not belong to the other family… It means that, that child should not stay with that family and the mother should take that child where it rightfully belongs*.*”* ***FGD 006***
>
> “*With results there are two things involved. If the results turn out to be good, this brings happiness, but if the results are not good, this will automatically bring violence in the house*.” **FGD 008**

### 2. Considerations for feedback of findings

Although respondents expressed willingness to receive feedback of genetics testing results, they highlighted several requirements that need to be put in place before results are shared.

#### Adequate Informed consent

Respondents observed the need for adequate informed consent before testing is carried out and at the time of feedback of results. Informed consent would facilitate individuals’ understanding of what they are getting involved in as well as the associated implications. This would help the individuals to make an informed decision about whether to be tested or not as well as their need for feedback of results. As highlighted below

> *“May be improving on learning resources or materials to make people understand things better*.*’ ‘* **FGD006**
>
> *“Availing information out there to the people explaining the importance of doing it and why we do it can really play a very big role in causing (a positive) change inattitude and perception of the people. So that is what I would suggest that we continue doing. It will be good to reach out to more people*.*’*’ **FGD006**
>
> ‘*‘If they come and teach me very well on what exactly they want to do plus let me know of the cons I can accept to participate in this DNA testing, it will also let me know my health status and they will also help me in case I have any health complications*.*’*’ **FGD 007**
>
> ‘*‘If they teach me well and I fully understand how this research works I can accept to participate*.*’ ‘* **FGD007**
>
> ‘*‘If they also tell me very well whereby, I also fully understand this research I can accept to participate so that I can be evidence to the community to let them know that it is not a bad initiative after all so that the research runs smoothly*.*’*’ **FGD007**

Respondents highlighted the need for research teams to facilitate participant understanding of the genetic information through a process which would include the use of visual aids in order to facilitate the information delivery process and promote understanding.

‘*‘What I think is that there should be a projector to show us photographic images of this genetic science and the procedure of genetic testing. When you see that the other child resembles the parents it makes you to appreciate that you are studying something that exists*.’’ **FGD 006**

Respondents highlighted the need for a clear appreciation of the condition being tested so that they are well informed of what is likely to happen to their bodies. They stressed the desire to know the results to the extent possible and if the information turns out to be complicated, then the feedback can involve their parents or close relatives. Thus, the need for the informed consent process to employ a number of visual aids to facilitate participant’ s understanding.

> *“If it were possible, I would want to see the nature of the disease through an image or explained to me thoroughly*.*’ ‘* **FGD 009**
>
> *‘’ I will accept because they would have taught me and I would have understood very well what they want to do. This will allow me to make up my mind and also I will know exactly what to do and also know what exactly is needed for my life*.*’ ‘* **FGD 007**

#### Genetic counselling

Adequate genetic counselling by a well-trained professional preferably a doctor was considered essential for individuals before getting their genetic testing results. Respondents felt that good counselling would help allay anxiety associated with receiving genetics results. Counselling would also help spell out any misconceptions or misunderstandings associated with genetics and genomics. Additionally, many respondents preferred to receive the feedback results in person because then the person giving the feedback of results would have an opportunity to guide them on which hospital can provide any necessary medication.

> “*Therefore, the counselling that is given before feedback of results from there (research centre) can excite and encourage me to continue to do that test and even to encourage my clan members on the same*.” **FGD 006**
>
> “*Of course, as doctors they know how to describe the results of a test; they should be professional in breaking the news to me*.” **FGD 009**
>
> “*Whoever is breaking the news has to sit calmly face to face with me to explain the results for me to understand well, without sending me into a shock*.*”* **FGD 010**

Respondents observed that before disclosure of the results and associated health condition, there should be a proper way of disclosing information and this may be by telling the individuals what to eat to prevent or control that condition. That It would be better to first advise the individuals on how to care for themselves and then disclose the condition. They also observed that It’ s not good to rush to disclose findings because someone might breakdown.

> *“The doctor should first counsel me because sometimes if they found a disease and they just gave me a paper, it can make me unsettled but if the doctor talks to me, tells me that we found a disease but take care of yourself, take your medication, this will make me not have any fears*.*’ ‘* **FGD 008**
>
> *“It is not right for a doctor to show my results to someone else to bring to me. This is because that person doesn’ t have the experience like the doctors who can counsel me*.” **FGD 007**
>
> *“For me I would need to be counselled properly before giving me the results*.*”* FGD010
>
> “*If my samples were picked from home, that means that my result should be brought home and also before giving me results they should counsel me but they should not just give me the results abruptly. My mind should be settled before giving me my results*.*”* **FGD 007**

#### Privacy and confidentiality

Respondents stressed the need for a quiet and private environment at the time of disclosing the results to individuals who have been tested. Since genetics and genomics testing results is regarded as private information, the need to observe privacy and confidentiality is a growing reality that should be respected at all times. Many respondents proposed that at the time of disclosing findings, it should be only the doctor and the individual who was tested.

> “*Earlier you talked about confidentiality which automatically means that in case they pick my sample for DNA my results will definitely be given to me meaning that whatever it is it will be between me, the doctors and the people carrying out the tests*.” FGD 007
>
> “*The results should be given to me directly. In case they find any medical complications, the person who has brought the results should explain to me their findings and also if possible, bring medicine and prescribe for me how to take the medication. If you give the results to someone else the person will begin telling people behind my back how my condition is very worrying and bad*.” **FGD 007**
>
> “*Whoever is breaking the news has to sit calmly face to face with me to explain the results for me to understand well, without going into a shock*.*”* **FGD 010**
>
> While most respondents felt that the results should be shared directly with the individual who was tested, some thought that they would need support and presence of a family member at the time of getting the results. Others proposed that if it’ s a condition that affects the family, then the doctor can disclose to the whole family. This would help everyone to know what condition is affecting the family since It’ s not something that can be kept secret.
>
> *“For me I want to be with my parents when I am getting the results from my hereditary testing*.*”* **FGD010**
>
> “*The results should be given to me personally because it is me supposed to tell my parents and also, I would want it in written form because the records can help me in future, let’ s* say *my condition becomes worse, I need to show those results at the hospital. If you get your results via message, you can’ t go and show a message to a doctor so I feel it’ s better to receive in written form*.” **FGD 007**

### 3. Extending feedback of genetics findings to family and community

#### Extending feedback to family

Respondents had varied opinions on extending feedback of results to their relatives with some stressing that the results only belong to the individual who was tested while others thought they could share findings with close family members. Some highlighted the fact that if one is likely to suffer from genetics related conditions, then it was necessary to share the findings of genetic testing with individuals who will take care of them in case they become sick.

> *“The family members need to know because some diseases may need extra attention and care like meals on time, special foods etc, so that the family members can be helpful in looking after you*.’’ **FGD010**
>
> “*I also feel it is right to tell my parents because it gives my brothers and also my wife the opportunity to also go and test in case, I turn out to be positive of any illness so that other children don’ t inherit the diseases too*.” **FGD 007**

Other reasons for extending feedback of genetics results to family included the need for informing others and help them know of their predisposition to disease early so as to take appropriate action. To others genetic information was considered a family health issue which affects all members of the family and so they have to be told the results.

> *“To me I think it depends on the type of disease because sometimes it might be a non-life-threatening condition or it can be like epilepsy which doesn’ t go hand in hand with noise so the people back home should know how to handle me. So, if the feedback of results is to me, then at least my parents have to be there and also, they should put in the records so that in case another disease comes in the doctors will have an idea on how to help me*.*’ ‘* ***FGD 007***
>
> *“It helps the family to understand the problem that they are faced with so that it’ s able to plan together, how to help in case there is any one sick and others are not, the family can understand how to plan and handle such situations*.*’* ***FGD 006***
>
> *“Because it helps on the side of treatment and health and unity of the family*.*’ ‘* ***FGD 006***

Reasons for not extending feedback of genetics testing results to family included the fear that some members may not understand the meaning of such information or would be unable to handle the associated stress and anxiety.

> *‘’ My view is that your test results should only be given to you because they are private and will only affect you*.*’ ‘* **FGD 010**
>
> “*These results will remain in my house; I will not share them out anyhow*.*”* **FGD 008**

#### Extending feedback of genetic results to the community members

Some respondents felt it was acceptable to share their genetics results with the community because they would support you in case you are unwell. Others observed that it was right to share because the DNA testing results do not necessarily mean that they are only testing for diseases. So trusted people in their circle who may not be relatives, can know the results and advise on what to do.

> *“For me I would tell all of them, so that they are aware of what the doctor has advised me to do and stand with me in support*.*’ ‘* **FGD010**
>
> *“It is good to share results with other people because it safeguards their health. If a person knows that I have a particular genetic disease, it will be up to them to decide whether to produce with me (children) or not. If a person chooses to marry me, that is their risk*.*’ ‘* ***FGD 006***
>
> *“It is good for the community to know because these days people assume someone’ s death up to the extent of accusing other community members with whom the person could have had a grudge, to avoid such assumptions, they should know*.*’ ‘* ***FGD010***
>
> *“I will accept that my wife should know, the community should also know, there are other diseases that can be spread, those near me can even help me if I am weak, my neighbours should also know*.*’ ‘* ***FGD 008***

Respondents who did not favour extending feedback of genetics results with community thought that it was private health information for the family that should not be shared with non-family members. Others felt like sharing such information exposes the health condition of the family and that might end up causing the family to be ridiculed or segregated.

“Yes, my results are important to my family members to know but not outside of family because sometimes that is a secret we have in our house.’’ **FGD 008**

*“It depends on the type of disease, if it is a disease that I can survive with by taking care of myself I feel it is ok to keep it to myself but if it is a condition that needs people’ s help like me getting lost, then the community should know about it*.’’ **FGD 007**

Some respondents who were opposed to extending feedback to the community felt like sharing such information might be used against you by some members of the community with resultant stigma and potential discrimination

> *“I think it’ s a bad idea because people who do not like you take advantage of the information to spread bad information about you and you become the talk of the town, so I think it’ s best to give it to the owner of the results*.*’ ‘* **FGD 010**

### Strategies for sharing feedback of genetics and genomics research results

Regarding the strategies for sharing feedback to family, several approaches were suggested by the respondents. Some respondents felt that they should have exclusive rights to disclose the information, hence the doctor should provide them with enough information that can be used to inform others. Some respondents thought the doctors would do the job of extending feedback of results to family members because they are better informed and equipped with the necessary genetics counselling skills. While others thought that they would pass information to an elder in the clan or family who would in turn take the responsibility of conveying the information to the rest of the family through approaches like family meetings.

“Alternatively, the testing team *can come home pick samples one by one (testing can be carried out at the home of the participants), it will be right to counsel me together with my parents so that they can know what to do, those are the ways our results can come back to us*.*”* **FGD 007**

## Discussion

We set out to assess the views of grassroots communities in Uganda on if and how feedback of hypothetical genetics and genomics research results can occur. Our study results show that this type of feedback of results was acceptable to all respondents. Several reasons for needing feedback of results were identified including and especially, the need to know one’ s health status and to plan for the future. Several strategies were proposed if such feedback was to be conducted appropriately.

The need to know one’ s health condition can be a benefit to research participants particularly in an African rural setting where genetic testing is out of reach of almost all individuals. Feedback to research participants is a growing reality and an ethical obligation that should be incorporated in the research processes as highlighted by several ethics guidelines [15, 29,30]. Although the usual call for feedback has been directed to other fields of research and just emerging in GGR because of the anticipated implications, such fears can be appropriately addressed via mechanisms like adequate consent processes and genetic counselling by qualified genetic counsellors, observance of privacy and confidentiality as well as sharing results that are potentially beneficial or actionable. Related work among genomics research participants and genomic researchers in Uganda has also highlighted the need for feedback of GGR results [27,28]. Additionally, need for feedback of GGR results has been considered by research participants in Botswana as a form of solidarity and as a reciprocity obligation of researchers that can make participants feel valued as part of a mutual relationship [21]. Dissemination, beneficence and reciprocity have been considered as essential components of a framework for enhancing ethical genomic research with indigenous communities in the USA [31]. Additionally, respondents in our study felt that knowing one’ s test results would help them seek early treatment or prevention, creating the impression that treatment for genetically predisposed conditions is available. Although this was a rural non-research setting, it’ s Important to note that therapeutic misconception where participants perceive research as care rather than experimentation are very common. And such misconception may mislead individuals into participating in research for diagnostic or therapeutic purposes, yet most genetic studies may not yield results that can benefit health or predict risk of disease. Even in cases where accurate diagnosis can occur, many diseases identified may not be treatable.

For feedback of genetics and genomics research results to be conducted appropriately, several strategies were proposed by the respondents including adequate consent processes, genetic counselling as well as privacy and confidentiality. Informed consent for research participants is an ethical requirement that should be carried out as a continuous process starting before recruitment, through to implementation in the post study period. Such consent processes should be suitable for participants and be provided in language that is easily understandable. The need for meaningful informed consent has been highlighted by participants of a genomics research study in Uganda that revealed recall bias about their participation in the concerned research study [27]. GGR has been challenged by the fact that genetics and genomics terminology and associated vocabulary may be difficult to translate in many of local languages in Uganda making it more difficult to achieve adequate and meaningful consent. Recent work that reviewed consent documents for 13 H3Africa genomics projects observed that genetics was mostly explained in terms of inherited characteristics, heredity and health, genes and disease causation, or disease susceptibility and only one project made provisions for the feedback of individual genetic results [30].Challenges regarding meaningful informed consent for GGR have been observed particularly when it comes to sharing of human biological samples and data in the context of international collaborative research [33, 34]. In order to address some of the challenges associated with informed consent in GGR, some commentators have proposed tailoring the informed consent process based on a ten-point framework which includes among others the study design, data and biological sample sharing, reporting study results to participants, cultural context, language and literacy and potential for stigmatization of study populations [35]. However, this proposed framework needs to be clearly interpreted and studied if it is to be meaningfully applied. In additional, for consent to be meaningful it should be coupled with relevant information on the proposed genetics testing and its implication. Such genetic counselling is essential and should be provided before testing and during feedback of results. Although genetic counselling is a developing field in emerging economies like South Africa [36], there is a relative lack of qualified genetic counsellors and the associated counselling in many of the low resource settings in Africa including Uganda [28,37]. Yet such genetic counselling would go a long way in addressing issues like implications of genetic testing and feedback of results to the individual, the family, the community, therapeutic misconception, privacy and confidentiality as well as the common beliefs in the Ugandan setting of genetic testing being primarily for paternity testing. Since the concept of genetic counselling is relatively new in our setting and virtually non-existent in the rural communities, respondents felt that the doctor who is most knowledgeable should be the one to conduct the counselling. This challenge can be addressed by capacity building for genetic counselling.

In addition, the consent forms should be explicit on aspects like who would have access to genetic results and whether return of results concerning paternity information should be done. If so, this should be approved by the REC before data collection is initiated. Otherwise, it’ s always a dilemma when researchers discover sensitive information after running the tests and seek guidance from an equally unprepared REC. For example, during genetic testing for sickle cell disease, which is prevalent in Uganda, it’ s not uncommon to discover discordant genetic information between the child and the male parent. It would be good if the consent documents approved by the REC clearly state if such paternity information will be provided to both parents.

Our study findings also highlight a situation where the participants stress the need for privacy and confidentiality of their genetic testing and return of results on one hand, yet most would prefer the presence of a family member during feedback of results a process which they thought could as well be done at participants’ homes. Hence the concept of confidentiality in these communities needs to be clarified and could imply keeping information not only to the individual tested but within their close family. Other aspects that need to be appropriately addressed to facilitate understanding of the genetics and genomics research concepts include meaningful community engagement (CE). Such engagement would help researchers understand community-based practices for example the meaning of privacy and confidentiality, and whether it should be handled at the individual level or family level. Some commentators have proposed the Tygerberg Research Ubuntu-Inspired Community Engagement Model which would require RECs/IRBs to play a role in requiring a CE plan for every study that is community based, and scientific journals to require a paragraph on CE in publication of relevant research projects. This would ensure moving CE from a guidance requirement to a regulatory requirement, emphasizing that it is a critical component of a robust consent process in research and that it ought to be embedded within research projects, where applicable [38].

Many respondents were agreeable to extending feedback of genetics and genomics testing results to family because genetics information was considered to belong to the whole family since it is inherited. The need for extending feedback to family and sometimes the wider community could be explained by the fact that most of the individuals in the Ugandan rural setting live in communities and support each other for their livelihood and during times of sickness. It is also important because family and community members play an important role in the provision of health care to patients. However, despite the fact that most of the respondents were agreeable to extending feedback of GGR results to family, fewer respondents supported such feedback to the wider community and only for particular health conditions. The practice of extending feedback of GGR results needs to be studied further and should be done on a case-by-case basis because the implications may vary across the different cases. This is in line with recommendations from a USA consultative team involving a working group of national experts of ELSI which among other recommendations suggested that researchers should elicit participants’ preferences on such extension of feedback to family but also recommended further research on the subject matter [39]. Other countries like Belgium have legislated laws that would allow health care professionals to disclose genetic information considered beneficial to family members in case the individual tested is not willing to do so [40].

It has also been recommended that it is imperative that the privacy and confidentiality of the person enrolling in the study should be respected but in cases where there is benefit in sharing results with family members, the original participant should grant permission because just like feedback to individuals, feedback should not be imposed on family members, but should be based on their voluntary consent [14]. A study involving REC chairpersons in the USA showed that 62% of the REC chairs agreed that participants should be informed that their results could be offered to family members and asked to indicate their choice, but such a statement may not be adequate informed consent [41]. Keeping genetic information and associated diseases confidential may be very difficult particularly in the Ugandan setting where the costs of medical treatment to a great extent are met by the relatives and sometimes the wider community who may inadvertently learn of the patients’ genetic condition. Since individuals in the communities are acceptable to extending feedback of GGR results to family members, it’ s up to the regulators to devise appropriate frameworks that would guide the process while respecting the participants’ preferences, privacy and confidentiality.

Finally, most GGR conducted in Uganda to address ethical, social and legal issues has been carried out in well-established research settings, and the views that have informed debate on ethical conduct of GGR in the country are mainly those of research participants, researchers and research regulators [25-28, 42]. Our addition of the grassroots communities will contribute a new dimension with an additional group of stakeholders whose views will enrich the literature as well as the proposed ethics guidelines for conduct of GGR in Uganda which was the goal of this study. We believe the ethics guidelines for conduct of GGR will go a long way in informing regulation and oversight of GGR in the country which is currently not guided by any specific guidelines.

### Limitations of the study

The individuals who participated in the study were research naive and may not have fully appreciated the implications of participation in GGR and feedback of the associated results.

Since the study was conducted in three different languages, the researchers needed assistance from individuals fluent in the respective languages to conduct the dFGDs and this might have affected the quality of the interviews and the subsequent data.

Additionally, the dFGD were conducted in three different local languages and later translated into English which could affect the quality of data.

Given the fact that this was a qualitative study, although the findings provide a deep understanding of the subject matter, they may not be generalizable. However, a wider range of other stakeholders have been involved in related research which enriches the generated data.

## Conclusion

Participation in hypothetical genetics and genomics research as well as feedback of testing results is acceptable to individuals in grassroots communities. While extending feedback of genetics and genomic research results to close family members was generally acceptable, extending feedback to the community was regarded as acceptable in a limited number of cases only. Reasons for needing feedback of genetics and genomics research results included the need to know one’ s health status and to plan for the future. The strong therapeutic misconception linked to GGR is concerning and has implications for consent processes and genetic counselling. Furthermore, the expectation of paternity testing results being embedded in all GGR needs to be managed appropriately. Privacy and confidentiality, benefits, risks as well as implications for sharing need to be considered for feedback of results to be conducted appropriately.

## Data Availability

The data underlying the results presented in the study are available and will be provided by the corresponding author if this manuscript is accepted for publication.

## Abbreviations

dFGDs: Deliberative Focus Group Discussions
GGR: Genetics and Genomics Research
REC: Research Ethics committee
UNCST: Uganda National Council for Science and Technology

## Availability of data and materials

Data sources are available on request.

## Authors Contribution

JO, BK, MM & KM conceptualized this study; JO, BK, JB, developed data collection tools; JO, BK & JB collected data; JO & BK analysed data; KM and MM reviewed all transcripts and edited all drafts of the manuscript. All authors provided substantive intellectual contributions to the study and manuscript and approve of its content.

## Acknowledgements

We are grateful to all the individuals in the various communities who participated in this study.

